# Higher long-term visit-to-visit blood pressure variability is associated with severe cerebral small vessel disease in the general population

**DOI:** 10.1101/2024.01.02.24300738

**Authors:** Xinyu Zhao, Hui Ying, Jing Li, Xianquan Shi, Shuohua Chen, Han Lv, Shouling Wu, Zhenchang Wang

## Abstract

**Background:** Long-term blood pressure (BP) variability is related to several diseases, but its impact on cerebral small vessel disease (cSVD) is uncertain. The study explored the relationship between BP variability, total cSVD burden, and its typical features.

**Method:** The study involved 1284 participants from the Kailuan cohort. From 2006 to 2022, systolic BP (SBP), diastolic BP (DBP), and pulse pressure (PP) variability were calculated as low, middle, or high. Magnetic resonance imaging was used to identify white matter hyperintensities (WMH), lacunae (LA), cerebral microbleeds (CMBs), and visible perivascular spaces (PVS). The burden of cSVD was defined as non, mild, moderate, or severe. Logistic regression was used to estimate the odds ratio (OR) and confidence interval (CI).

**Results:** High SBP variability was associated with moderate cSVD burden (OR=1.89, 95% CI: 1.09-3.29) and PVS (OR= 1.62, 95%CI: 1.10-2.39). The DBP was associated with LA (OR=1.74, 95%CI: 1.06-2.84). The PP showed obvious risk effects on moderate/severe cSVD burden (OR= 1.99, 95%CI: 1.17-3.41; OR=2.49, 1.34-4.63). These associations were modified by age and hypertension status. In the young adults (age<60 years old), only high PP variability associated with severe cSVD burden (OR=3.33, 95%CI: 1.31-8.44), LA (OR=3.02, 95%CI: 1.31-6.93), and PVS (OR=1.86, 95%CI: 1.20-2.88). The risk effect of SBP and PP variability on cSVD burden was only significant in the participants with hypertension.

**Conclusion:** High long-term BP variability, especially in combination with hypertension, is a risk factor for total cSVD burden, LA, and PVS. it is crucial to pay attention to PP variability in young adults.

## 1 Introduction

Cerebral small vessel disease (cSVD) is a chronic progressive disorder that refers to a group of conditions affecting the small blood vessels in the brain ^1^. Typical identified cSVD subtypes include white matter hyperintensities (WMH), lacunae of presumed vascular origin (LA), cerebral microbleeds (CMBs), and visible perivascular spaces (PVS)^2^. Patients with cSVD are at an increased risk of stroke, transient ischemic attack depression, and vascular dementia, finally causing disability and mortality^3^. The disease’s mechanism remains unclear, leading to no practical preventative or therapeutic approach for cSVD^4^. Total cSVD burden, which sums up the different magnetic resonance imaging (MRI) features that were present, was a simple and effective indicator that captured the overall effect of SVD on the brain. It is meaningful to explore factors that increase the risk of incidence of cSVD or the severity of cSVD burden, finding potential measures for disease prevention.

The concept of visit-to-visit blood pressure (BP) variability is not new^5^. Numeric evidence has suggested that BP variability is a significant independent risk factor for several diseases, especially cardiovascular events, regardless of average BP^6–8^. Though a meta-analysis showed that BP variability was associated with cSVD, the positive relationship was only significant using 24-hour ambulatory BP monitoring and short-term (<6 months) visit-to-visit BPV^9^. The evidence on the relationship between long-term visit-to-visit BP and cSVD is still limited. Furthermore, most existing studies assessed WMH, while evidence on the total cSVD burden and other cSVD subtypes, including LA, CMBs, and PVS, is sparse.

So, based on the Kailuan cohort study, the present study aimed to explore the relationship between long-term visit-to-visit BPV and total cSVD burden and every typical cSVD feature.

## 2 Method

### 2.1 Study population

This is a cross sectional study based on an ongoing longitudinal prospective cohort study, the Kailuan study, as detailed elsewhere ^10,11^. From 2006 to 2007, 101 510 participants in the Kailuan community were enrolled, and follow-up surveys were conducted biennially. The study has finished eight surveys (2006-2007, 2008-2009, 2010-2011, 2012-2013, 2014-2015, 2016-2017, 2018-2019, and 2020-2022). At the recent survey (2020-2022), we additionally performed a multi-modality medical imaging study (META-KLS), which enrolled subjects who participated in the Kailuan study and were asked to complete multi-modality medical imaging, including brain MR imagingscan, retinal fundus photograph, and so on. The detailed rationale and design of META-KLS has been published online^12^. The study enrolled 1508 participants who agreed on the META-KLS and completed brain MR imaging. We excluded 224 participants due to the following: (1) having less than three visits of BP measure during the past eight surveys (N=174); (2) having a stroke at present examination (N=3); (3) having cancer at present examination (N=1); and (4) more than half of the covariate information is missing (N=46). Finally, 1284 participants were included in the analysis (Figure 1.).

**Figure 1.**
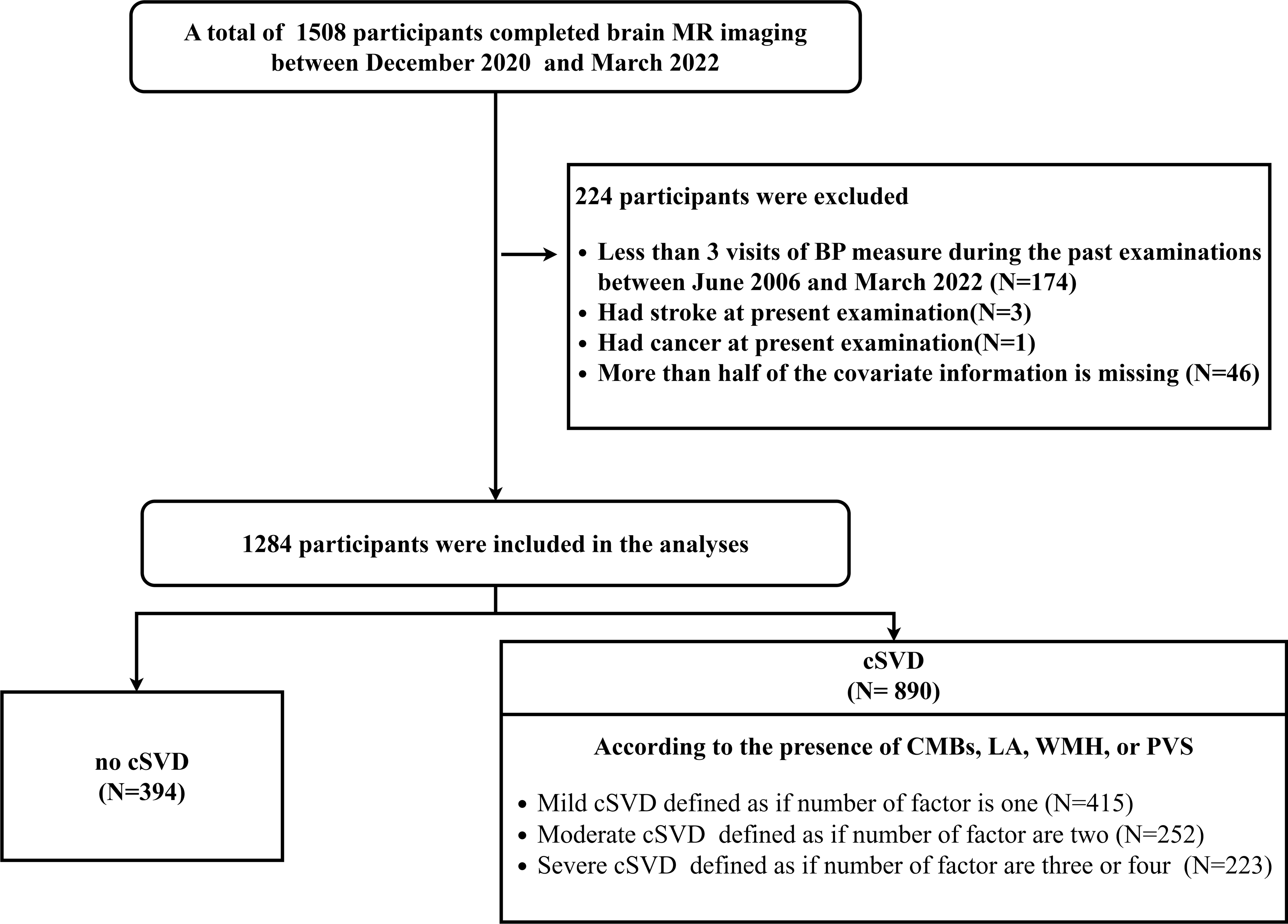
The flow chart of the study participants. **Abbreviations:** Cerebral small vessel disease (cSVD) white matter hyperintensities (WMHs), lacunae of presumed vascular origin (LA), cerebral microbleeds (CMBs), visible perivascular spaces (PVS), magnetic resonance (MR), and blood pressure (BP).

### 2.2 Ethics Statement

The Medical Ethics Committee of Kailuan General Hospital has approved both the Kailuan Study and META-KLS (IRB Number: 2008 No.1 and 2021002, respectively). These two cohort studies were registered online (ChiCTR2000029767 on chictr.org.cn and NCT05453877 on Clinicaltrials.gov, respectively). Informed consent was obtained from all participants included in the study.

### 2.3 Variability of BP assessment

The BP, including systolic BP (SBP) and diastolic BP (DBP), was measured in the left upper arm using a calibrated mercury sphygmomanometer while the participant sat. At least 2 BP measurements were taken after 5 min of rest. BP was then measured again if the difference between the two measurements was more than 5 mm Hg. The final BP value was recorded as the average value of the BP measurements, and pulse pressure (PP) was calculated as SBP minus DBP. Hypertension was defined as systolic blood pressure ≥140 mm Hg and diastolic blood pressure ≥90 mm Hg, currently receiving antihypertensive treatment, or having a self-reported physician diagnosis.

Four indices of variability were used: (1) coefficient of variation (CV), (2) standard deviation (SD), and (3) variability independent of the mean (VIM); VIM was calculated as SD/mean. It is similar to CV except that the mean BP denominator is raised to a specific power, x, that removes any correlation with mean BP. Power x is modeled as SD = k × means and was derived from fitting curves by nonlinear regression analysis as implemented in the PROC NLIN procedure of the SAS package^5^. (4) average natural variability (ARV) is the average absolute difference between successive values^13^. The VIM was used as the primary variability measure.

### 2.4 MR imaging and definition of cSVD

The brain MR imaging was collected using a 3.0 Tesla scanner and an eight-channel phased-array coil (GE 750W; General Electric Medical Systems, Milwaukee, WI, USA). The sequences used in the MRI examination included the following: axial T2-weighted imaging, three-dimensional (3D) brain volume (BRAVO) for high-resolution T1-weighted imaging, 3D fluid-attenuated inversion recovery imaging, diffusion-weighted imaging (DWI), and susceptibility-weighted angiography (SWAN).

According to the Standards for Reporting Vascular Changes on Neuroimaging 2 (STRIVE-2), ^2^,4 closely correlated features are markers of cSVD were assessed, including (1) LA, which was defined as the presence of one or more lacunes (1 point if present); (2) CMBs, which was defined as the presence of one or more CMB (1 point if present); (3). PVS, which was defined as there were moderate to severe (grade 2–4) PVS in the basal ganglia (1 point if present); (4) WMH, which was defined as either (early) confluent deep WMH (Fazekas score 2 or 3) or irregular periventricular WMH extending into the deep white matter (Fazekas score 3) (1 point if present). The total burden of cSVD was rated on an ordinal scale from 0 to 4 by counting the presence of each of the 4 MRI features of cSVD. And then defined as a categorical variable, including non-cSVD if the total burden is zero, mild cSVD if the entire burden is one, moderate cSVD if the total burden is 2, and severe cSVD if the total burden is 3 or 4.

All images, including WMH, Lacunes, CMBs, and PVS, were independently evaluated by two experienced neuroradiologists. The consistency of vision has been proved in our previous study^14^.

### 2.5 Potential confounders

We conducted questionnaire surveys to collect the following information, including age (18-59 years old, or ≥60 years old), gender, education level (junior high school or below, senior high school, or college and above), smoking status (never smoking, past smoking, or current smoking), drinking status (never drinking, past drinking, or everyday drinking), physical activity, disease history, and medications. We categorized the physical activity level as “inactive,” “moderately active,” or “active” according to the frequency of physical activity (>20 min/time) during leisure time; if the response was “never,” “sometimes,” or “≥ four times/week,” correspondingly. Trained field workers measured height and weight, and body mass index (BMI) was calculated as weight in kilograms divided by height in meters squared. Laboratory tests, including fasting blood glucose, triglycerides (TG), high-density lipoprotein cholesterol (HDL-c), and low-density lipoprotein cholesterol (LDL-c) were assessed in the central lab. Diabetes was defined as fasting blood glucose ≥7.0 mmol/L, taking oral hypoglycaemic agents or insulin, or having a self-reported physician diagnosis.

### 2.6 Statistical methods

Variables were presented as mean ± standard deviation or number (%) and compared using analysis of variance or Chi-square test, respectively.

For the test of parallel lines was not fitted, multinomial logistic regressions were used to estimate odds ratios (ORs) and 95% confidence intervals (CIs) of the BP variability on the total burden of cSVD (mild *vs.* non, moderate *vs.* non, and severely non). Unconditional logistic regressions assessed the association between BP variability and the four markers of cSVD (WMH, LA, CMBs, and PVS). BP variability was categorized into three groups (< p25 as low, p25-p75 as middle, or ≥ p75 as high). Three multivariate models were constructed to control for possible confounding effects: (1) age- and gender-adjusted model, which adjusted baseline age (2020-2022 survey) and gender; (2) multivariate model 1, which further adjusted education level, drinking status, smoking status, physical activity level, baseline BMI, diabetes, TG, HDL, and LDL level. (3) Multivariate model 2 is based on multivariate model 1 and further adjusted the mean value of SBP, DBP, or PP when the exposure was the corresponding variability of the BP indicator.

Assuming that missing data was random, we used multiple imputations by chained equations to impute one complete dataset with five interactions, which showed equal distribution before and after imputation (Supplementary Table 1). Subgroup analyses were performed according to age group, gender, and hypertension status to test the potential modification effect. We also conducted sensitivity analyses to verify the results: (1) using the original complete dataset, (2) excluding participants with diabetes, (3) excluding participants who are currently taking antihypertensive medications.

The main results were shown using the multivariate model 2. For the detailed results, see the supplementary tables 3-10. Analyses were implemented with SAS version 9.4 (SAS Institute Inc. Cary, NC) or R version 3.5.2 (https://www.r-project.org/).

## 3 Results

### 3.1 Baseline characteristics and the prevalence of cSVD burden

Of the invited 1284 participants, the mean age was 56.5±11.8 years old, and 52.7% were male. The mean follow-up duration was 13.1±3.0 years. cSVD was identified in 890 (69.3%) participants at the baseline, including 212 (16.5%) LA, 351 (27.3%) CMBs, 785 (61.1%) PVS, and 346 (27.0%) WMH. For the patients with cSVD, 415(46.6%) had mild cSVD burden, 252(28.3%) had moderate cSVD burden, and 223(25.1%) had severe cSVD burden (supplementary table S2). Compared with the participants without a cSVD burden, the subjects with a cSVD burden were significantly older, were more male, had lower education but higher physical activity levels, were generally smokers and drinkers, were usually diabetes, hypertension, had slightly higher BMI and LDL-c level, while had lower HDL-c level. There was no significant difference between the participants with and without cSVD in follow-up durations. (table 1)

**Table1.** Baseline characteristics of subjects with different levels of cSVD.

| Variables | All<br>(N=1284) | non-cSVD<br>(N=394) | mild cSVD<br>(N=415) | moderate cSVD<br>(N=252) | severe cSVD<br>(N=223) | *p* value |
| --- | --- | --- | --- | --- | --- | --- |
| Age (n, %) |  |  |  |  |  |  |
| 18-59 years old | 795(61.9) | 368(93.4) | 280(67.5) | 98(38.9) | 49(22.0) | <0.001 |
| ≥60 years old | 489(38.1) | 26(6.6) | 135(32.5) | 154(61.1) | 174(78.0) |  |
| Male (n, %) | 678(52.8) | 156(39.6) | 198(47.7) | 154(61.1) | 170(76.2) | <0.001 |
| Follow-up duration, years (mean±SD) | 13.1±3.0 | 13.0±2.8 | 13.1±3.1 | 13.0±3.2 | 13.3±2.8 | 0.691 |
| Education level (%) |  |  |  |  |  | <0.001 |
| Junior high school or below | 521(40.6) | 88(22.3) | 153(36.9) | 138(54.8) | 142(63.7) |  |
| Senior high school | 258(20.1) | 81(20.6) | 90(21.7) | 49(19.4) | 38(17.0) |  |
| College and above | 505(39.3) | 225(57.1) | 172(41.4) | 65(25.8) | 43(19.3) |  |
| Physical activity level (n, %) |  |  |  |  |  |  |
| Inactive | 173(13.5) | 68(17.3) | 54(13.0) | 26(10.3) | 25(11.2) | <0.001 |
| Moderate | 972(75.7) | 302(76.6) | 319(76.9) | 187(74.2) | 164(73.5) |  |
| Frequently | 139(10.8) | 24(6.1) | 42(10.1) | 39(15.5) | 34(15.3) |  |
| Smoking status (n, %) |  |  |  |  |  |  |
| Never | 921(71.7) | 316(80.2) | 311(75.0) | 167(66.3) | 127(56.9) | <0.001 |
| Past | 51(4.0) | 11(2.8) | 12(2.9) | 16(6.3) | 12(5.4) |  |
| Current | 312(24.3) | 67(17.0) | 92(22.1) | 69(27.4) | 84(37.7) |  |
| Drinking status (n, %) |  |  |  |  |  |  |
| Never | 727(56.6) | 237(60.1) | 248(59.8) | 142(56.3) | 100(44.8) | <0.001 |
| Past | 20(1.6) | 3(0.8) | 3(0.7) | 4(1.6) | 10(4.5) |  |
| Current | 537(41.8) | 154(39.1) | 164(39.5) | 106(42.1) | 113(50.7) |  |
| Diabetes (n, %) | 199(15.5) | 33(8.4) | 47(11.3) | 58(23.0) | 61(27.4) | <0.001 |
| Hypertension (n, %) | 724(56.4) | 138(35.0) | 219(52.8) | 179(71.0) | 188(84.3) | <0.001 |
| BMI, kg/m<sup>2</sup> (mean±SD) | 25.3±3.5 | 25.1±3.7 | 25.2±3.5 | 25.7±3.2 | 25.7±3.4 | 0.048 |
| TG, mmol/L [median(p25,p75)] | 1.3(2.6,3.7) | 1.3(0.9,2.0) | 1.4(1.0,2.0) | 1.4(1.0,2.1) | 1.4(1.0,2.0) | 0.875 |
| HDL-c, mmol/L (mean±SD) | 1.5±0.4 | 1.5±0.4 | 1.5±0.4 | 1.4±0.3 | 1.4±0.4 | <0.001 |
| LDL-c, mmol/L (mean±SD) | 3.2±0.8 | 3.1±0.8 | 3.3±0.8 | 3.2±0.8 | 3.1±0.9 | 0.001 |

Abbreviations: cSVD: cerebral small vessel disease; non-cSVD: if the total burden is zero; mild cSVD: if the total burden is one; moderate cSVD: if the total burden is 2; and severe cSVD: if the total burden is 3 or 4; BMI: body mass index; TG: triglycerides; HDL-c: high-density lipoprotein cholesterol; LDL-c: low-density lipoprotein cholesterol.

### 3.2 Associations between BP variability and burden of cSVD/different markers of cSVD

Figure 2 shows the associations between BP variability and the burden of cSVD. Compared with low variability of SBP, higher SBP variability was significantly associated with moderate cSVD burden (middle *vs* low: OR=1.66, 95%CI: 1.03-2.65, *p* = 0.036; high *vs* low: OR=1.89, 95%CI: 1.09-3.29, *p* = 0.023). High SBP variability showed a risk effect on the severe cSVD burden on the boundary of significance (OR=1.75, 95%CI: 1.09-3.29, *p* = 0.067). For the markers of cSVD, High SBP variability was significantly associated with PVS (OR= 1.62, 95%CI: 1.10-2.39, *p* = 0.015). High DBP variability was only observed to be associated with LA (OR=1.74, 95%CI: 1.06-2.84, *p* = 0.082). High PP variability showed more obvious risk effects on moderate/severe cSVD burden (moderate cSVD burden: OR= 1.99, 95%CI: 1.17-3.41, *p* = 0.012, severe cSVD burden: OR=2.49, 1.34-4.63, *p* = 0.004), LA (OR=1.75, 95%CI:1.06-2.89, *p* = 0.028), and PVS (OR=1.77, 95%CI: 1.20-2.61, *p* = 0.004).

**Figure 2.**
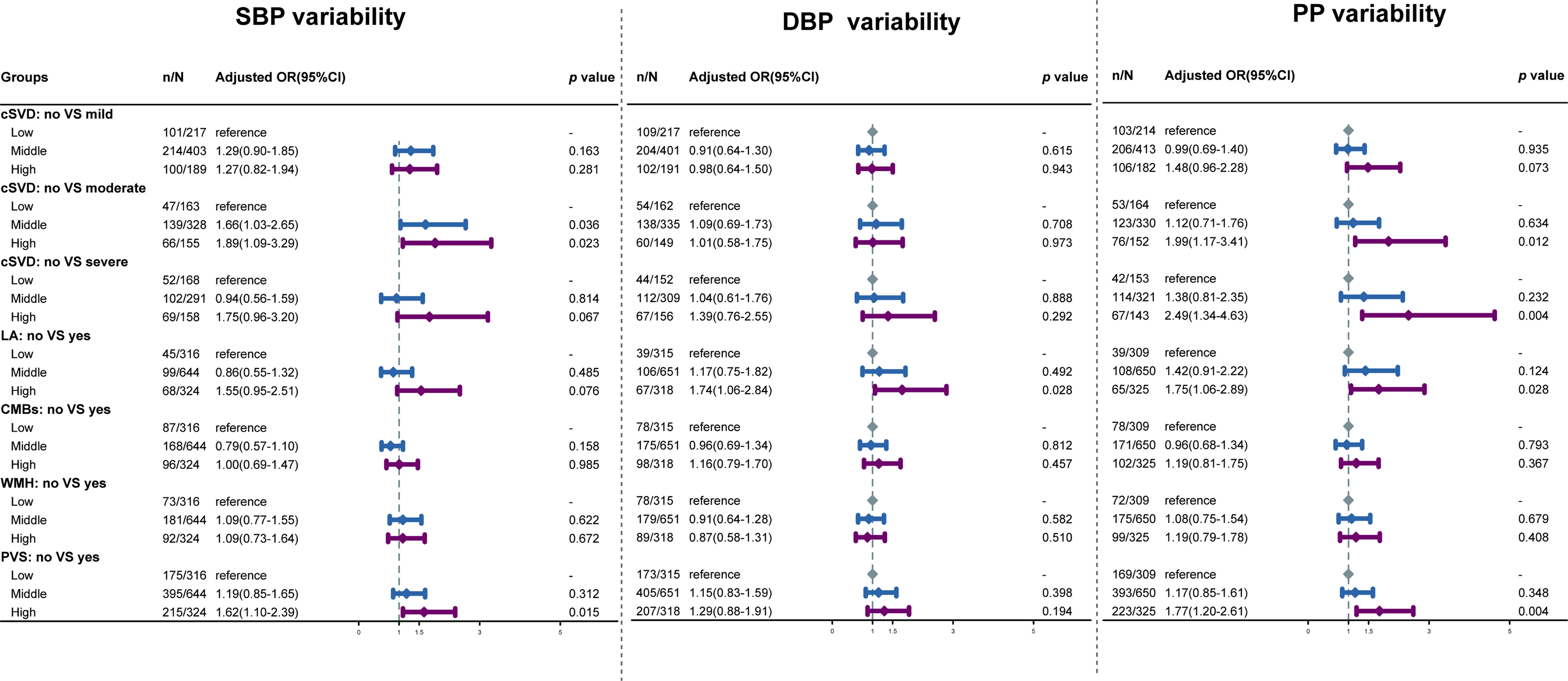
The associations between BP variability, total cSVD burden, and typical features of cSVD. **Abbreviations:** Cerebral small vessel disease (cSVD) white matter hyperintensities (WMHs), lacunae of presumed vascular origin (LA), cerebral microbleeds (CMBs), and visible perivascular spaces (PVS), systolic blood pressure (SBP), diastolic blood pressure (DBP), and pulse pressure (PP). Adjusted OR: Adjusted for baseline age (2020-2022 survey), gender, education level, drinking status, smoking status, physical activity level, baseline BMI, diabetes, TG, HDL, LDL level and the mean value of SBP, DBP, or PP when the exposure was the corresponding variability of the BP indicator.

### 3.3 Associations between BP variability and burden of cSVD/different markers of cSVD by age groups

Figure 3 shows the estimated risk of BP variability on cSVD burden and different markers of cSVD by age group. For participants under 60 years old, high variability of SBP was significantly associated with PVS(OR=1.65,95%CI: 1.06-2.58, *p* = 0.027), High variability of PP was significantly associated with moderate (OR=2.04,95%CI: 1.03-4.06, *p* = 0.042), severe cSVD burden (OR=3.33,95%CI: 1.31-8.44, *p* = 0.011), LA (OR=3.02,95%CI: 1.31-6.93, *p* = 0.009), and PVS(OR=1.86,95%CI: 1.20-2.88, *p* = 0.006). The variability of DBP was not observed to be associated with LA for participants 60 years or older (OR=2.37,95%CI: 1.26-4.45, *p* = 0.007). Only high variability of SBP showed a significant risk effect on the moderate cSVD burden (OR=4.87, 95%CI:1.14-20.83, *p* = 0.033) and severe cSVD burden on the boundary of significance (OR=3.87, 95%CI: 0.90-16.74, *p* = 0.070).

**Figure 3.**
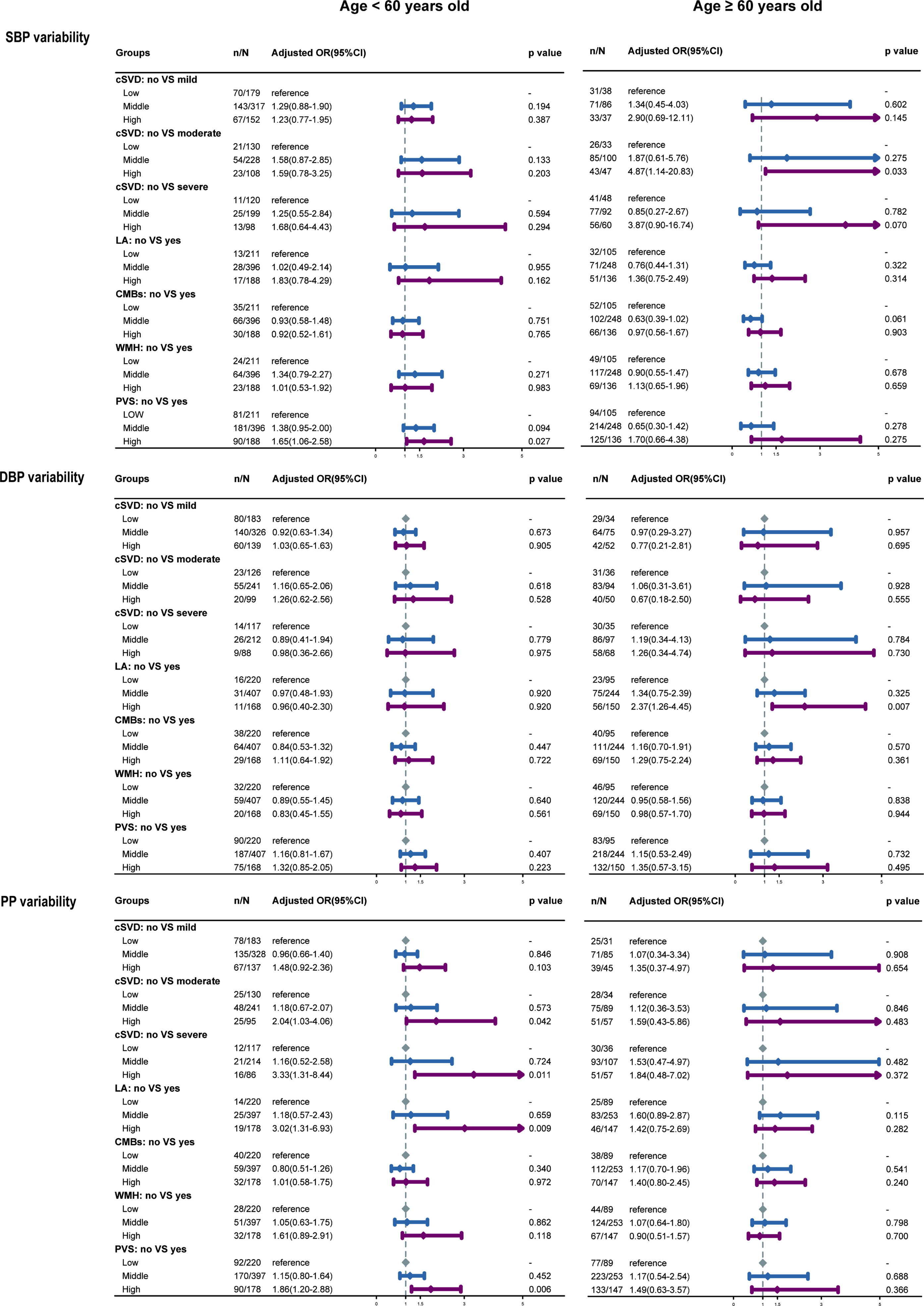
The associations between BP variability, total cSVD burden, and typical features of cSVD by age groups. **Abbreviations:** Cerebral small vessel disease (cSVD) white matter hyperintensities (WMHs), lacunae of presumed vascular origin (LA), cerebral microbleeds (CMBs), and visible perivascular spaces (PVS), systolic blood pressure (SBP), diastolic blood pressure (DBP), and pulse pressure (PP). Adjusted OR: Adjusted for gender, education level, drinking status, smoking status, physical activity level, baseline BMI, diabetes, TG, HDL, LDL level and the mean value of SBP, DBP, or PP when the exposure was the corresponding variability of the BP indicator.

### 3.4 Associations between BP variability and burden of cSVD/different markers of cSVD by gender and hypertension status

The association between BP variability and cSVD burden or different markers of cSVD were consistent both in males and females (Figure 4). Stratified by hypertension status, the risk effect of higher variability of SBP (middle vs low variability for moderate cSVD burden: OR=2.10, 95%CI:1.05-4.18, *p* = 0.035) and PP (high vs low variability for severe cSVD burden: OR=2.32, 95%CI:1.03-5.22, *p* = 0.042) on cSVD burden were only significantly positive in the participants with hypertension. (Figure 5).

**Figure 4.**
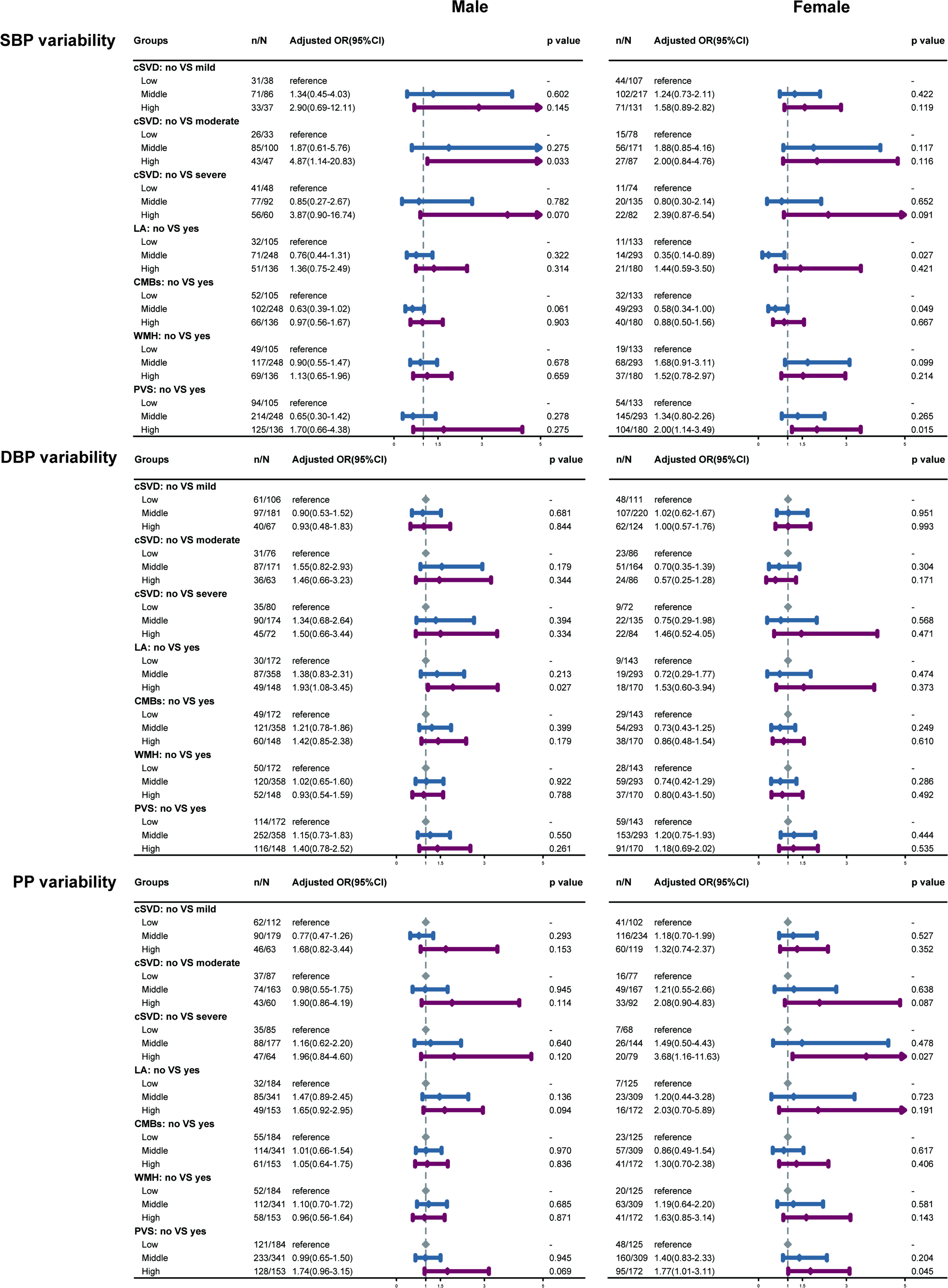
The associations between BP variability, total cSVD burden, and typical features of cSVD by gender. **Abbreviations:** Cerebral small vessel disease (cSVD) white matter hyperintensities (WMHs), lacunae of presumed vascular origin (LA), cerebral microbleeds (CMBs), and visible perivascular spaces (PVS), systolic blood pressure (SBP), diastolic blood pressure (DBP), and pulse pressure (PP). Adjusted OR: Adjusted for baseline age (2020-2022 survey), education level, drinking status, smoking status, physical activity level, baseline BMI, diabetes, TG, HDL, LDL level and the mean value of SBP, DBP, or PP when the exposure was the corresponding variability of the BP indicator.

**Figure 5.**
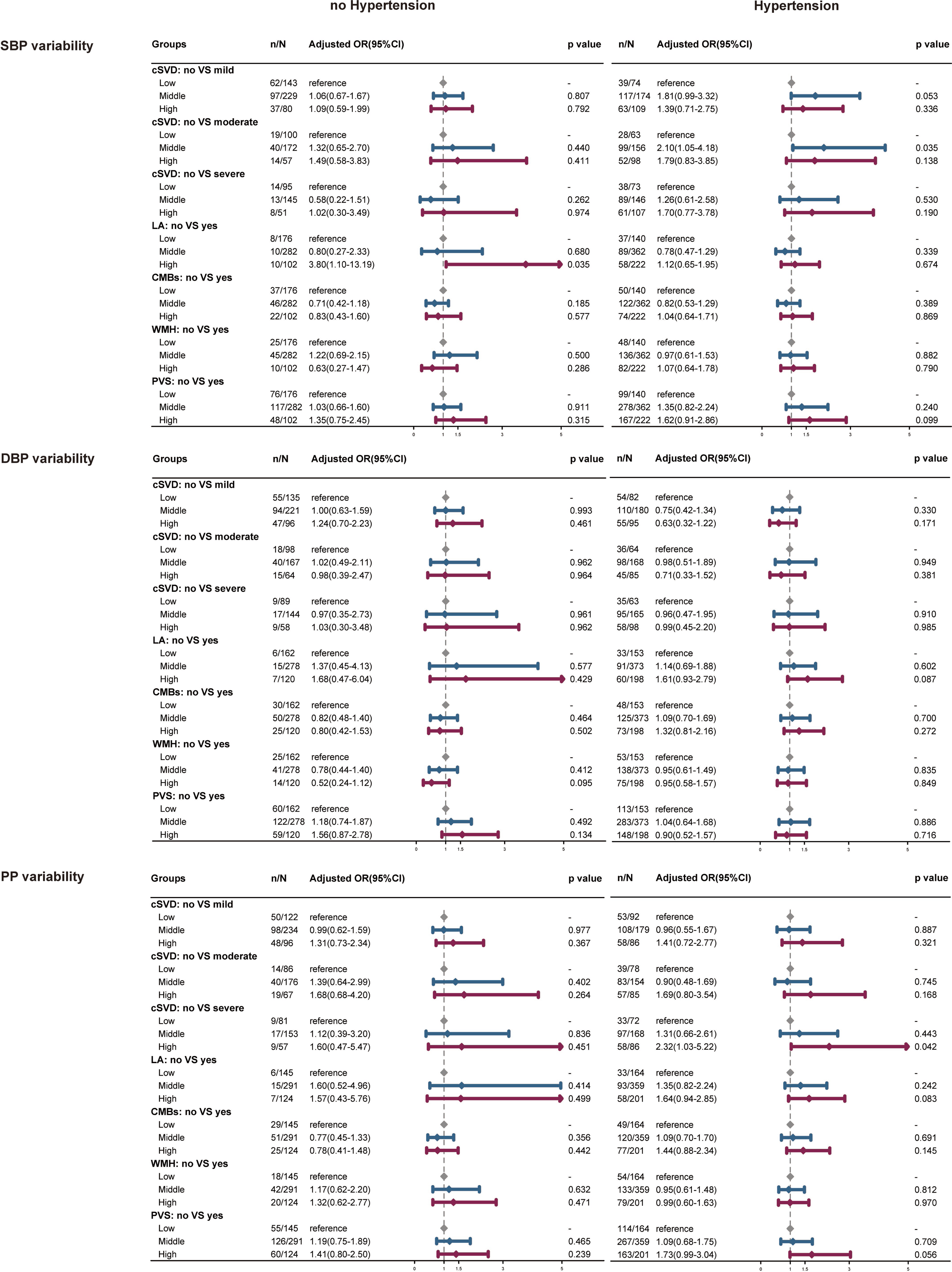
The associations between BP variability, total cSVD burden, and typical features of cSVD by hypertension status. **Abbreviations:** Cerebral small vessel disease (cSVD) white matter hyperintensities (WMHs), lacunae of presumed vascular origin (LA), cerebral microbleeds (CMBs), and visible perivascular spaces (PVS), systolic blood pressure (SBP), diastolic blood pressure (DBP), and pulse pressure (PP). Adjusted OR: Adjusted for baseline age (2020-2022 survey), education level, drinking status, smoking status, physical activity level, baseline BMI, diabetes, TG, HDL, LDL level and the mean value of SBP, DBP, or PP when the exposure was the corresponding variability of the BP indicator.

### 3.5 Sensitivity analyses

Using original data, excluding the participants with diabetes, excluding participants who are currently taking antihypertensive medications, or using other variability indicators, the estimated impact of BP variability on cSVD burden and different markers did not change substantially (table 2, supplementary table 11-12).

**Table2.**
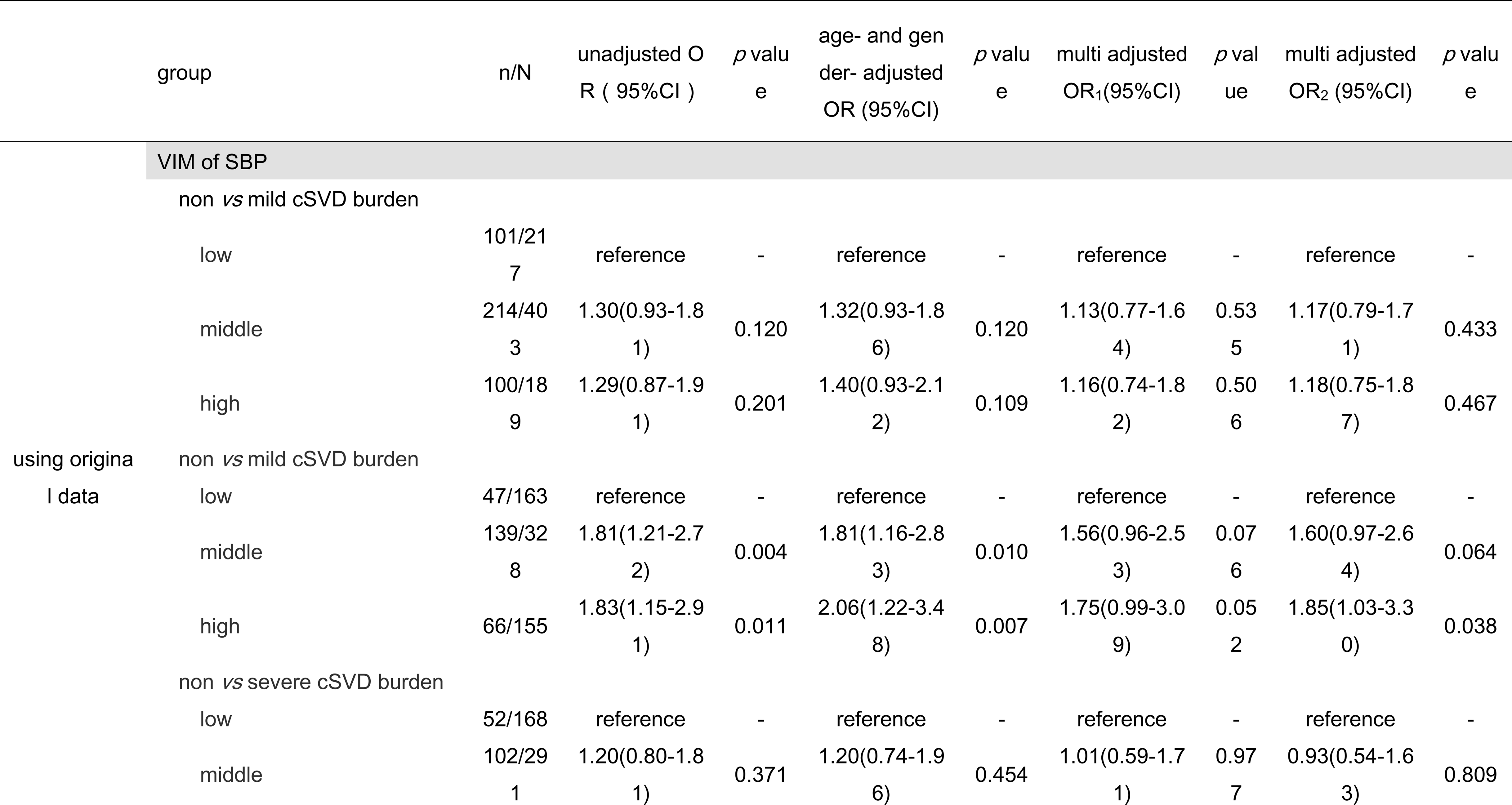
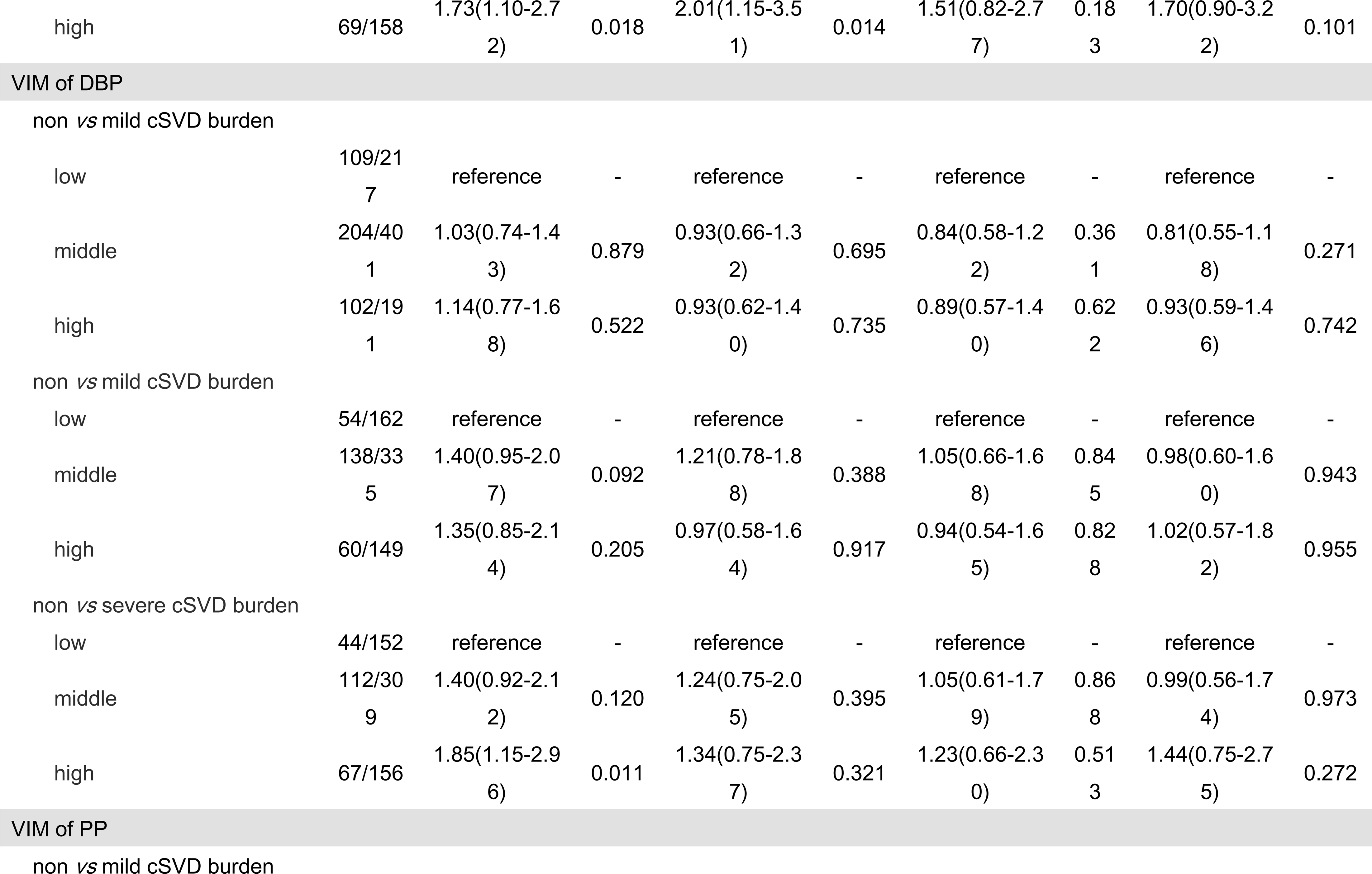
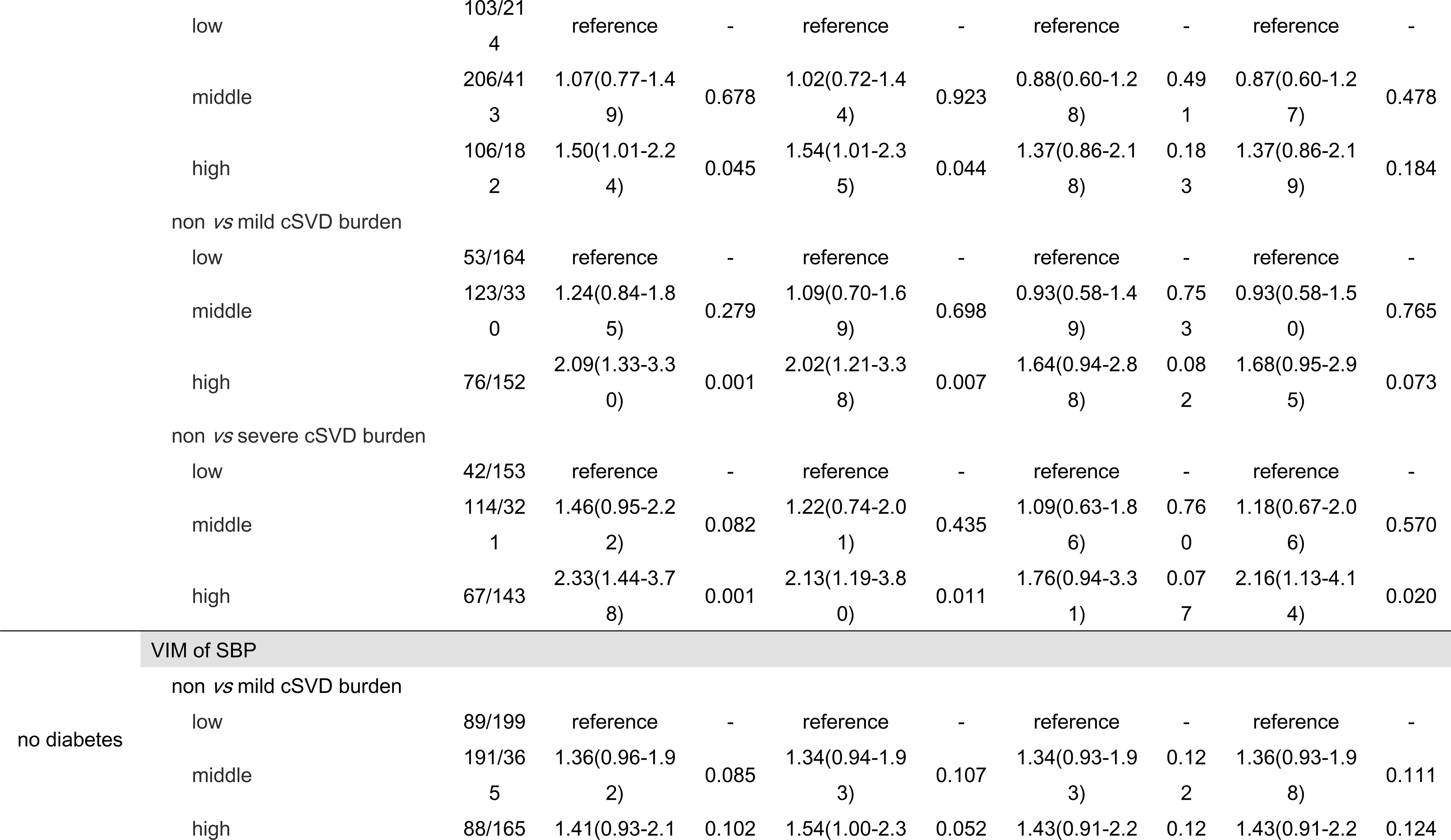
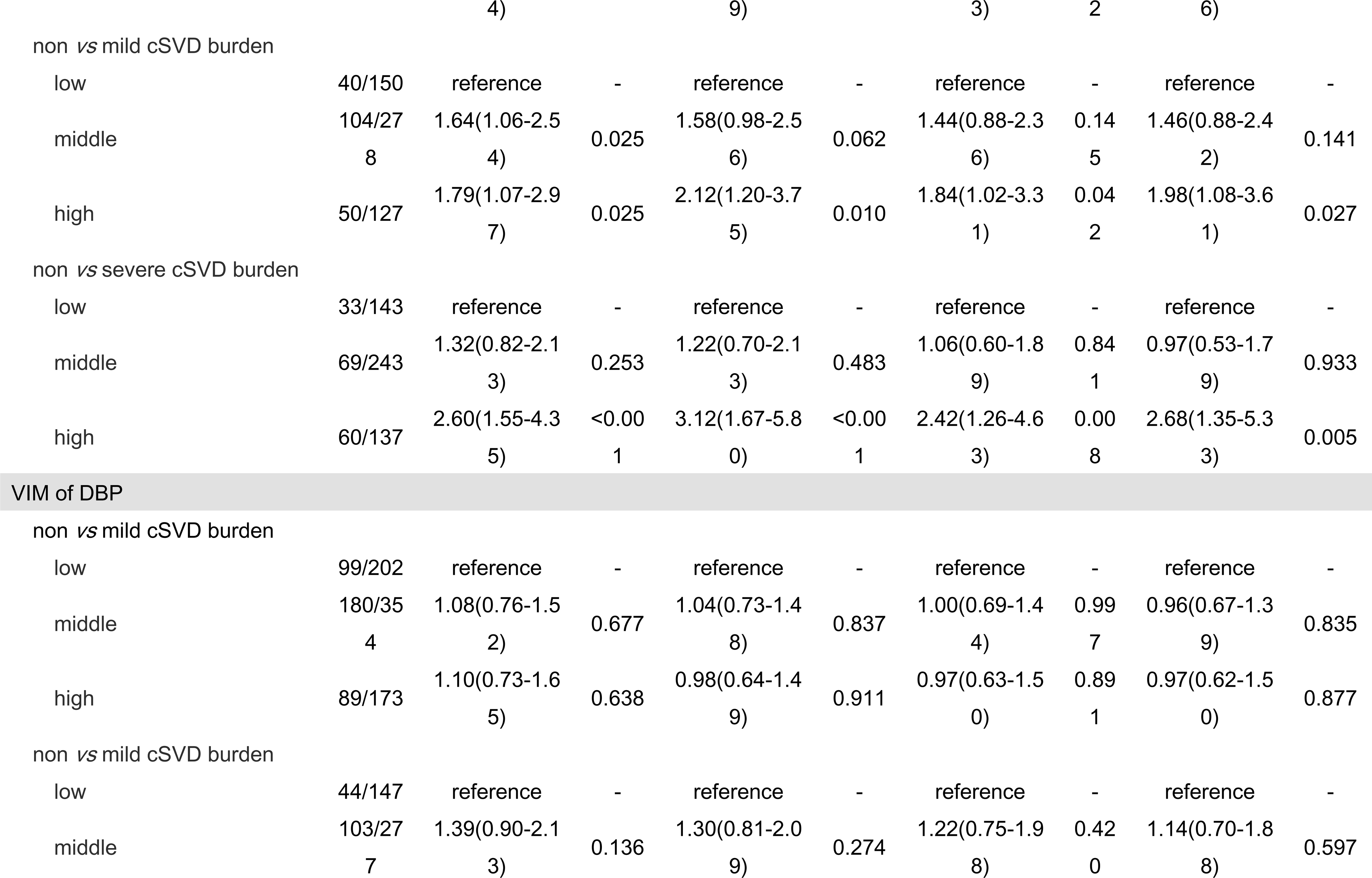
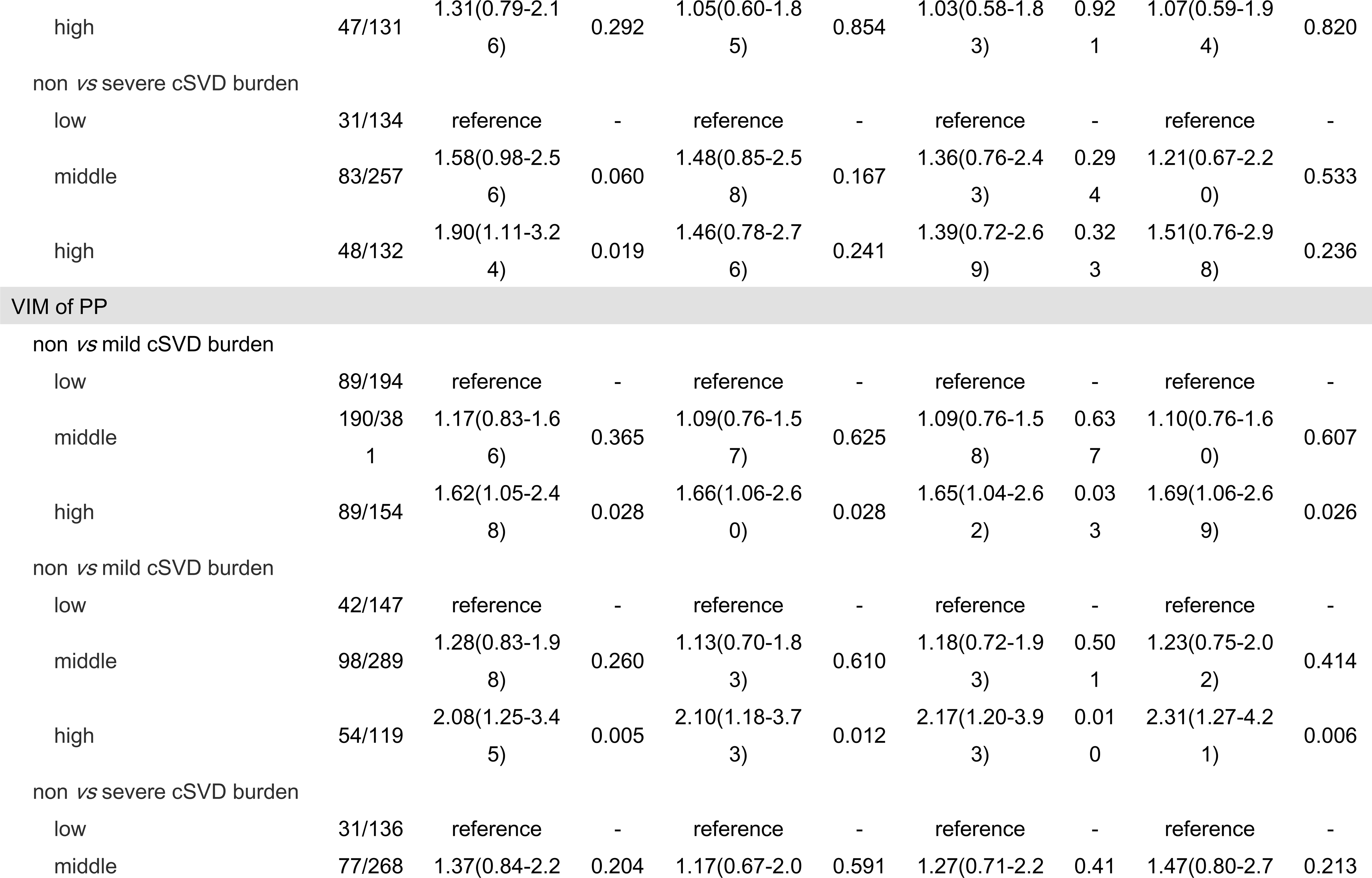
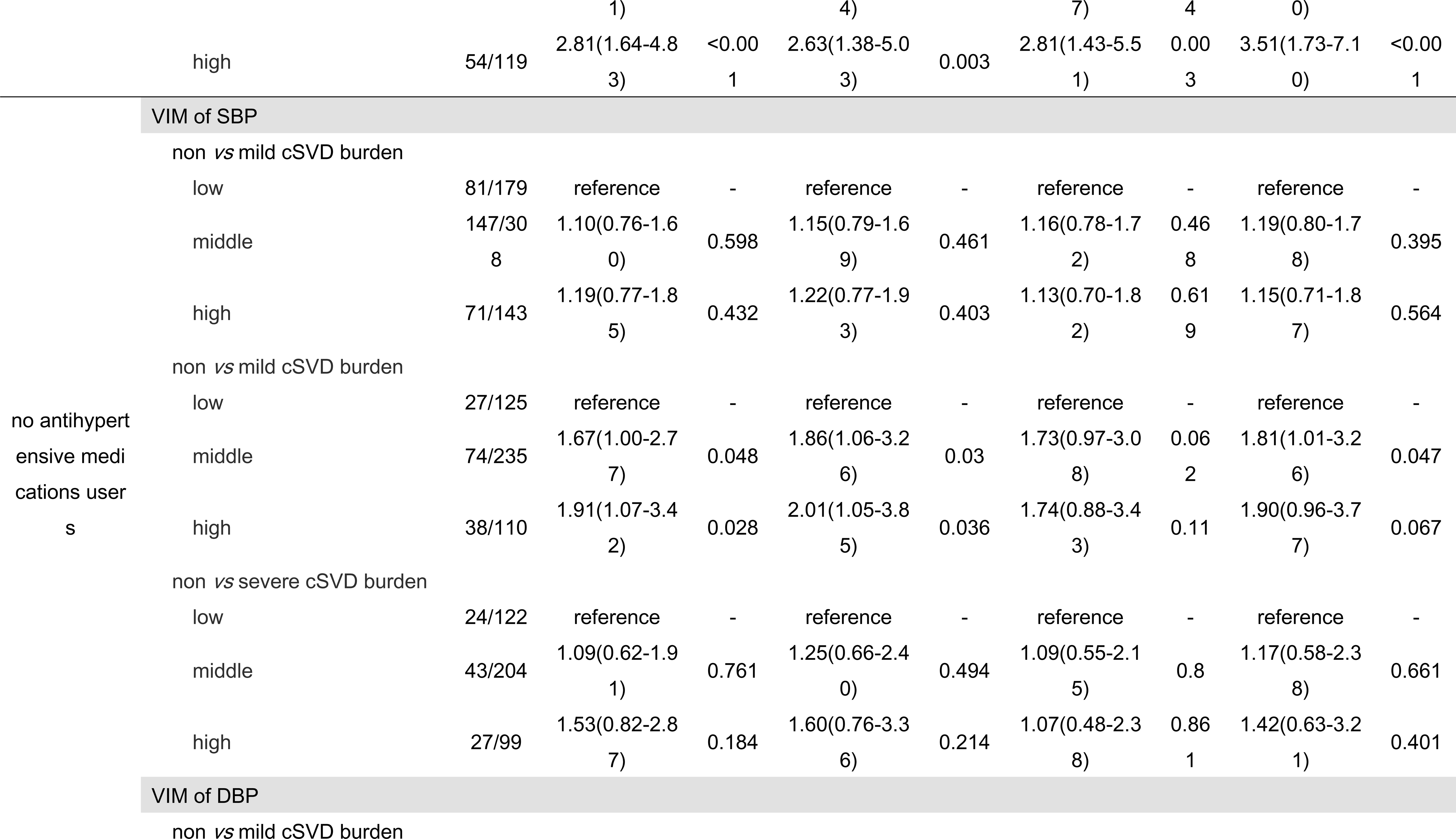
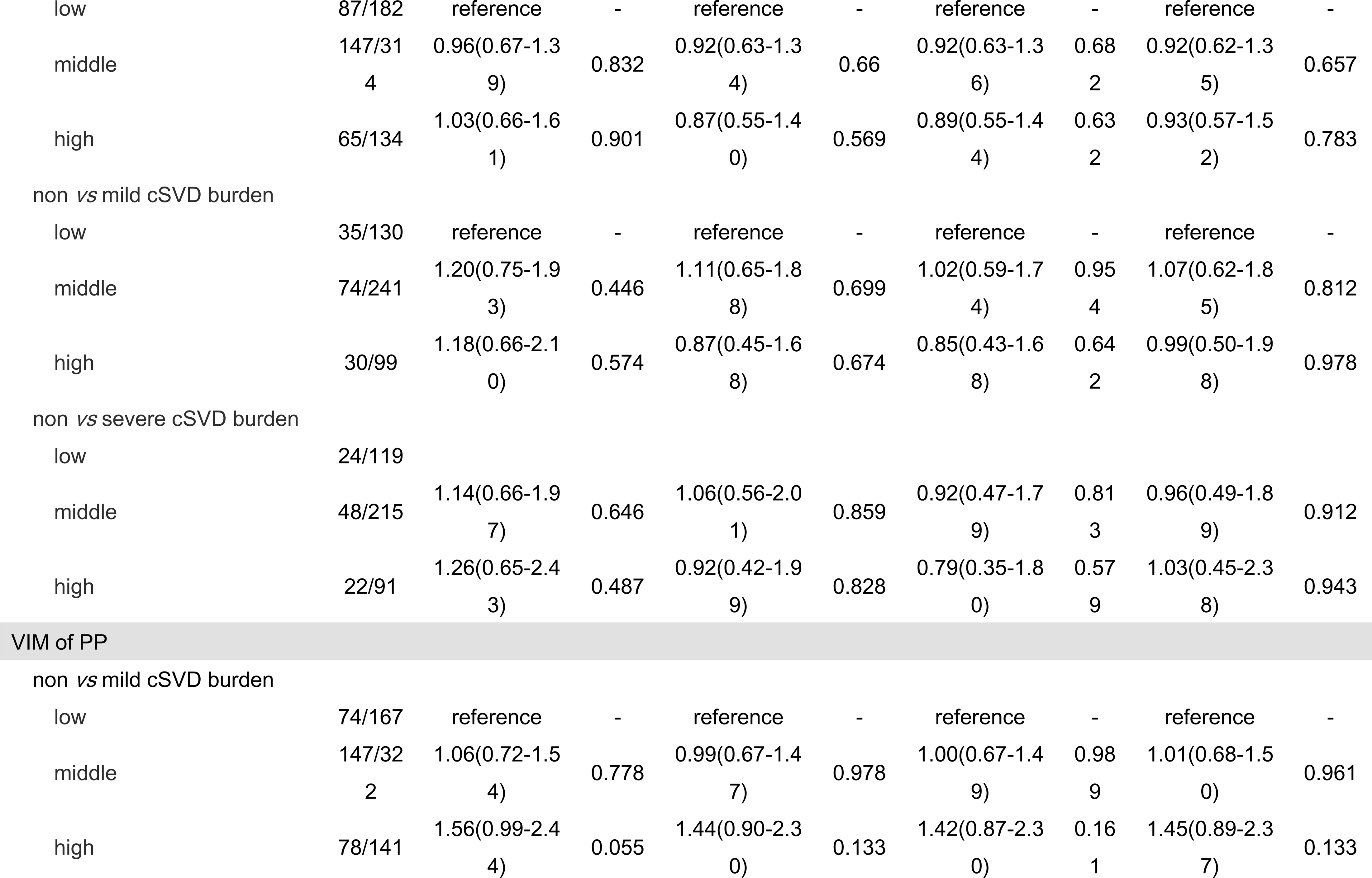
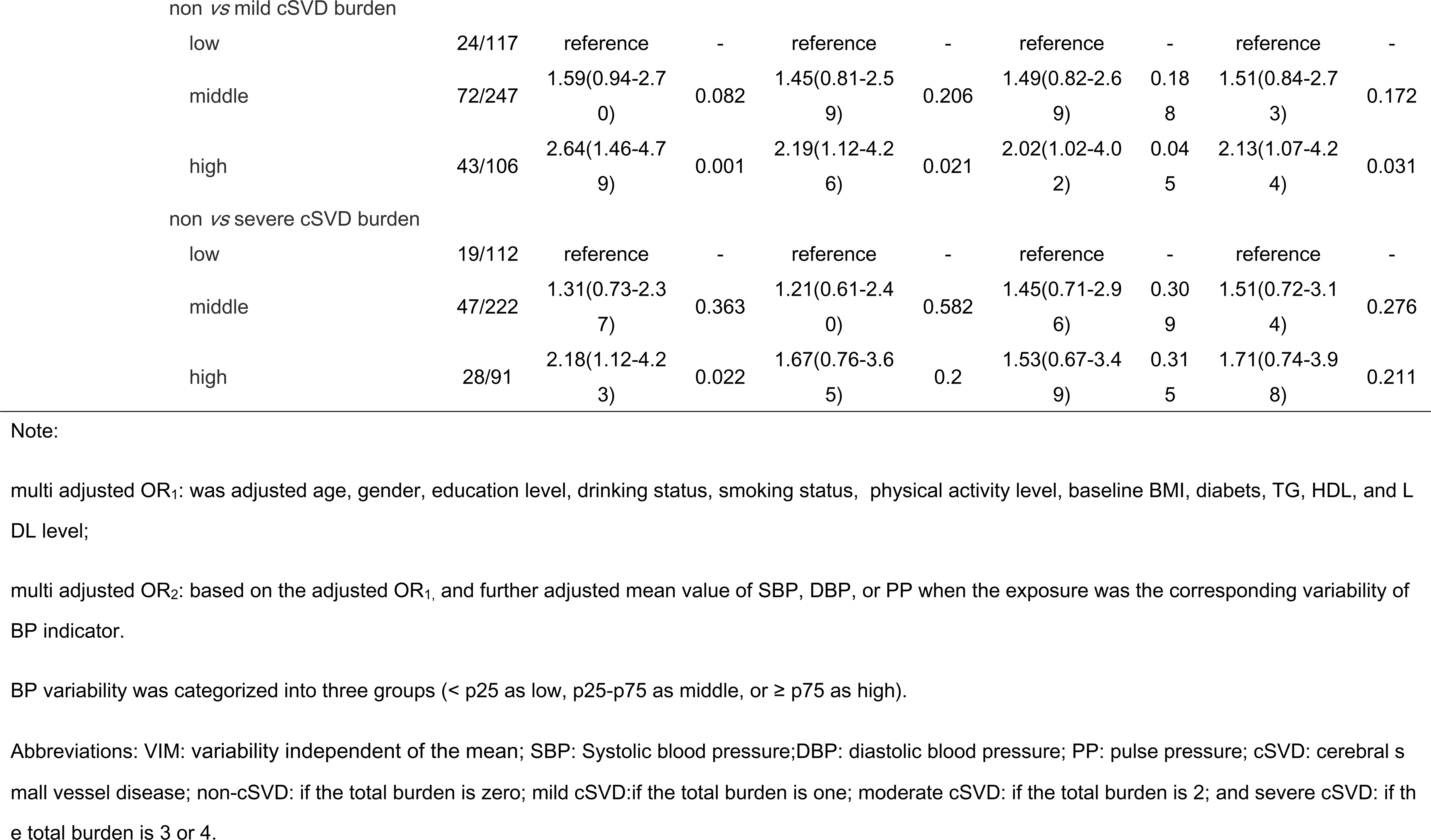
Sensitivity analysis for the estimated association between variability of BP and cSVD burden.

## 4 Discussion

In this study, we identified cSVD in 890 (69.3%) participants in this sub-study of a population-based cohort. We showed that higher long-term visit-to-visit BPV was an independent risk factor for cSVD, including the total cSVD burden, LA, and PVS. Notably, we found that the risk effect of BPV on cSVD differs by age.

Our study showed that higher visit-to-visit variability in SBP, not DBP, was significant with total cSVD burden. Recently, two meta-analysis studies have demonstrated that BPV was independently associated with CSVD^9,15^. However, these studies primarily defined the cSVD as WMH. Until now, little study has explored the relationship between long-term visit-to-visit BPV and total cSVD burden. Two other studies showed that short-term (twenty-four-hour)^16^ or middle-term (day-to-day)^17^ SBP variability was associated with total cSVD burden but did not observe a significant association between DBP variability and total cSVD burden, which was consistent with our study to a certain extent. The underlying mechanisms between BPV and cSVD are still poorly understood. There are several possible explanations. Firstly, high BPV may increase blood flow pulsation and dampen blood flow smoothing, causing damage to brain microvasculature^18^. Secondly, high BPV can potentially hinder nitric oxide production and compromise endothelial function. This could lead to injuries in the “neurovascular unit,” abnormalities in the blood-brain barrier, and, consequently, lesions in small blood vessels^19–21^. It has been proposed that increased variability in blood pressure may stem from pre-existing brain diseases disrupting central autonomic control^22^. Calcium channel blockers, recognized as the most effective drug class for mitigating variability in blood pressure, have demonstrated noteworthy efficacy in reducing the risk of vascular cognitive impairment^23,24^. This observation underscores the potential clinical significance of agents targeting blood pressure variability in diminishing the risk of brain vascular disease and cognitive impairment in old age.

For the specific markers of cSVD, we found that the risk effect of long-term variability on SBP was mainly on the PVS (high *vs* low: OR= 1.62, 95%CI: 1.10-2.39, p = 0.015), and DBP variability only had a risk effect on LA (high *vs.* low: OR=1.74, 95%CI: 1.06-2.84, *p* = 0.082. Studies have shown that the high level of office SBP, 24-hour ambulatory SBP^25^, and cumulative SBP were the risk factors for PVS^26^. Our study showed that the long-term variability of SBP was also an independent risk factor for PVS. The relationship between DBP and LA has not been thoroughly studied yet. Furthermore, our study showed that long-term variability of PP seemed to have a more significant risk effect on total cSVD burden, LA, and PVS than SBP and DBP. Previous studies evaluated BPV most frequently using SBP or DBP, few studies employed PP. So far, studies have reported that PP was a better predictor of cardiovascular^27^ and cerebrovascular^28^ risk than diastolic or systolic pressure. Studies showed PP was associated with the cognitive independence of SBP^29,30^. Increased PP and its variability might represent diminished regulatory functions of vessels against pulsate blood flow, making the brain tissues more susceptible to direct injury.

We did not observe a significant association between BPV and other markers of cSVD, including WMH and CMBs. However, most existing studies showed that BPV was a risk factor for WMH. These studies mostly used the SD CV as the BPV indicator but did not consider the potential confounding effect of the mean value of BP^31–34^. It is difficult to determine whether BPV has an independent risk effect on cSVD besides the absolute BP level. Our data also showed that high SBP variability of SD was significantly associated with WMH (OR=1.73, 95%CI:1.15-2.62, p=0.009). However, this association disappeared when the mean level of SBP was adjusted, which was consistent with a previous study. It demonstrated that the BPV is closely related to the absolute level of BP and may act on some common pathways for WMH^35^. The relationship between BPV and CMBs remains limited and conflicting ^15,35–37^, which need more research to identify their relationship.

An interesting finding in our present study is that the risk effect of BPV on cSVD differs by age. High variability of PP significantly affected total cSVD burden, LA, and PVS only for patients aged < 60 years old. Higher PP is related to cerebral microvascular damage; an animal experiment recently showed that its risk effect is age-dependent^38^. The greater stiffness of cerebral arteries from old potentially protects against the negative consequences of high PP variability. It suggested that more attention for PP besides SBP and DBP for young adults is needed to reduce the risk of cSVD. For patients aged ≥ 60 years old, the risk effect of high SBP variability on total cSVD burden and high DBP variability on LA appeared. It showed that SBP variability has an overall risk effect on cerebral small vessel disease, rather than by considering only one individual feature separately. Additionally, our data suggested that the risk effect of BP variability may modified by hypertension status. The risk effect of high BP variability was more evident in the participants with hypertension. It may be due to the hypertension group was more likely to have high fluctuation in blood pressure, for poor medication compliance. So, BP management, including both the high absolute level and variability of BP, is crucial for hypertension patients to prevent cSVD.

For the strengths, previous studies mainly focused on some specific features of cSVD, especially for WMH, while evidence on LA and PVS is sparse. The present study explored the relationship between BPV and four features of cSVD, including LA, WMH, CMBs, and PVS, in a relatively large sample. Furthermore, we verified the overall risk effects of BPV on the total cSVD burden, which takes all features of cSVD. Novelly, we found that the risk effect of BPV on cSVD differs by age. For young adults (age<60), steady fluctuation of PP was also an important intervention target, which needs more research to confirm. From a methodological standpoint, we employ VIM as the primary measure of variability. VIM was explicitly developed as a new measure of variability uncorrelated with mean blood pressure^5^. In our multivariate model, we adjust the mean blood pressure level, indicating that BPV impacts cSVD independently of mean blood pressure. Several limitations of our study should be noted. Firstly, MRI examinations were only performed once in the current survey, so potential reverse causation remains a major issue. This study is a nested case-control study derived from a 13-year follow-up cohort, ensuring the measurement accuracy for exposure, confounders, and outcome. Additionally, our study is still in progress, and the population with no cSVD will still be followed up. The relationship between BPV and the occurrence of cSVD will be retested in future studies. Secondly, the participants receiving the MRI were a small proportion of the original cohort; there were still likely potential selection biases beyond the confounders included in the study.

## Conclusions

Higher long-term visit-to-visit BP variability was an independent risk factor for cSVD, including the total cSVD burden, LA, and PVS, especially for the participants with hypertension. For young adults (age<60 years old), higher PP variability may be a more critical risk factor for cSVD than SBP and DBP.

## Acknowledgment

The authors thank all the members of the Kailuan Study Team for their contribution and the participants who contributed their data.

## Data Availability

Data are available on reasonable request from the corresponding author at.

## Abbreviations

(cSVD): Cerebral small vessel disease
(WMHs): white matter hyperintensities
(LA): lacunae of presumed vascular origin
(CMBs): cerebral microbleeds
(PVS): visible perivascular spaces
(MR): magnetic resonance
(BP): blood pressure
(SBP): systolic blood pressure
(DBP): diastolic blood pressure
(PP): pulse pressure
(META-KLS): multi-modality medical imaging study

---

**Table 3.**
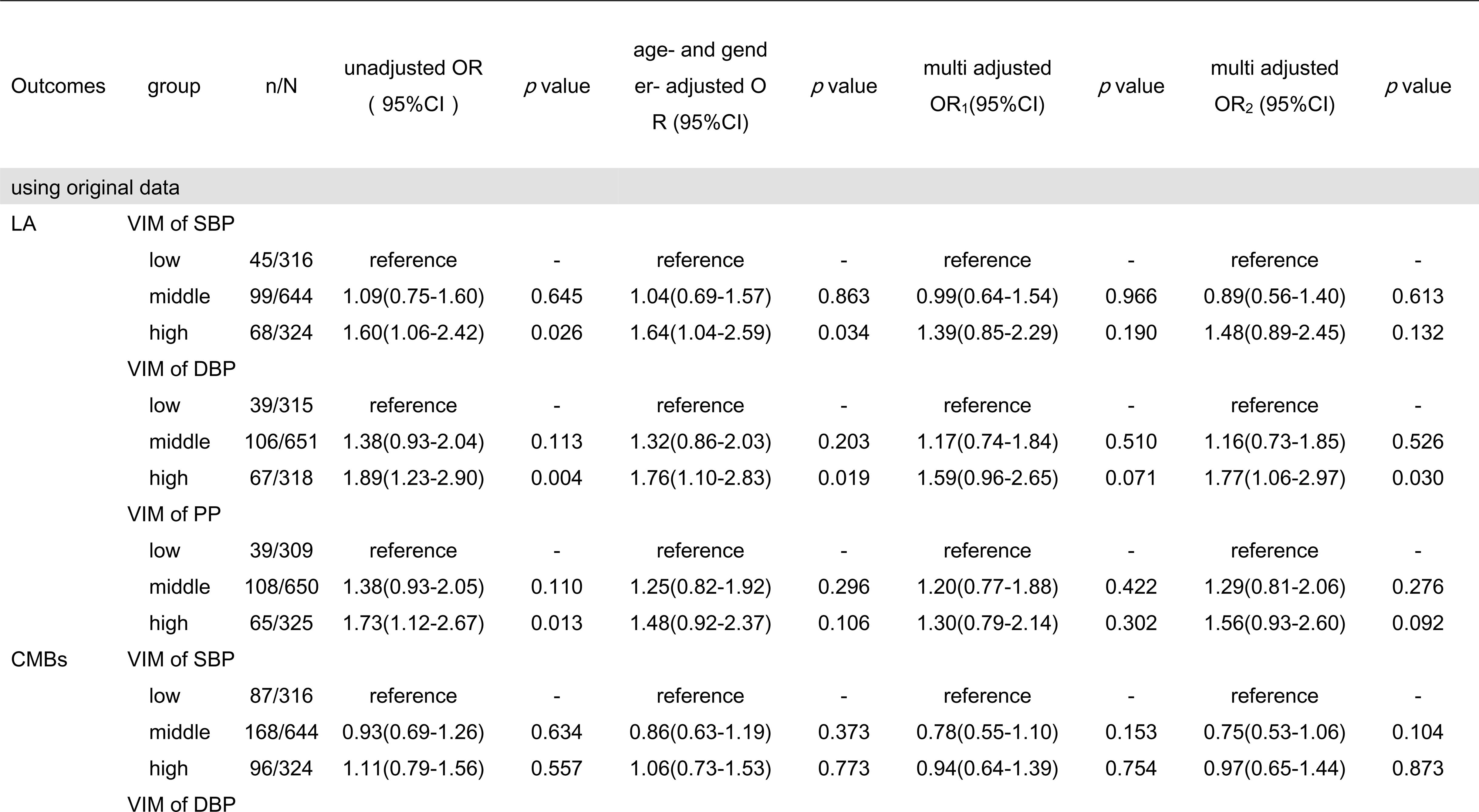
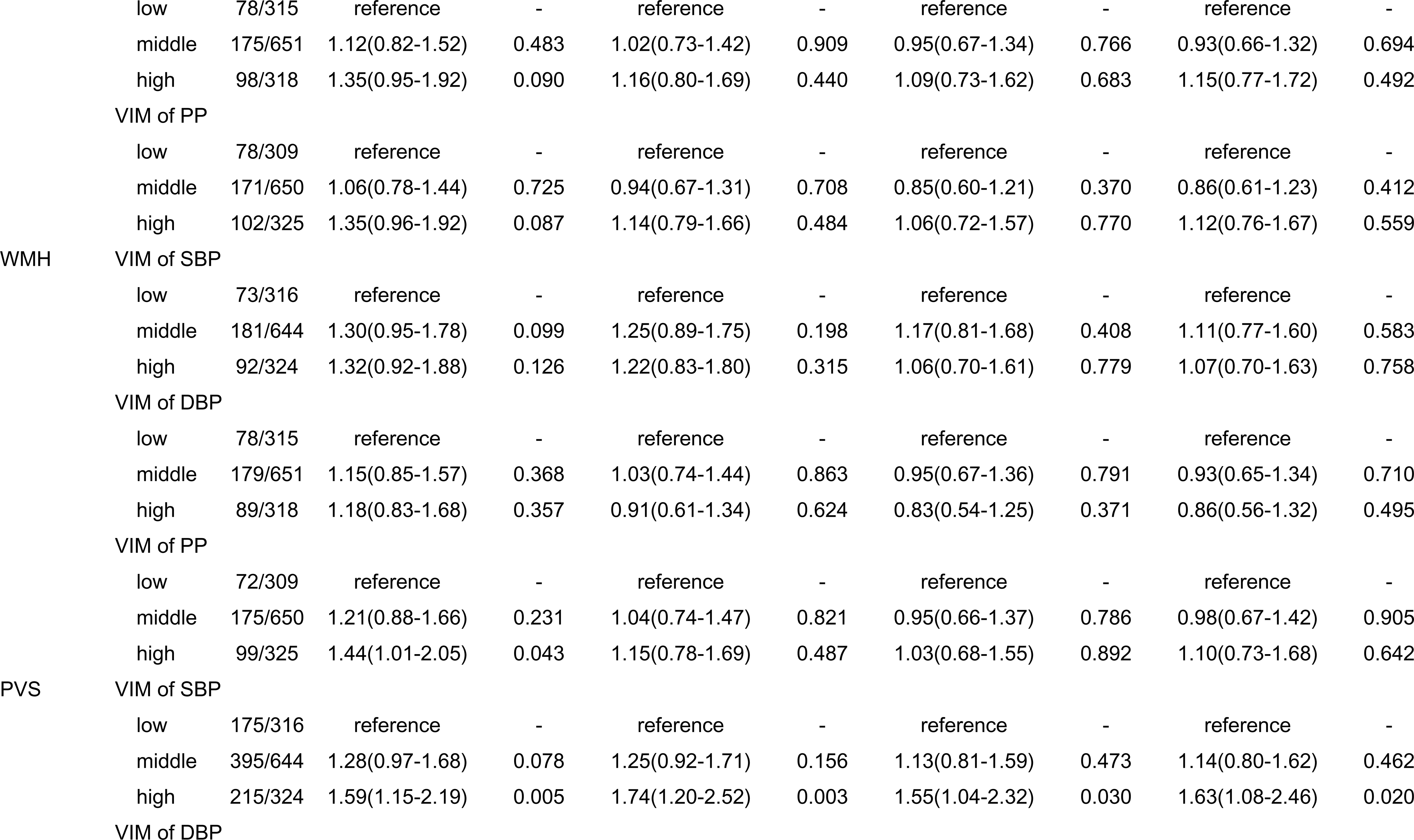
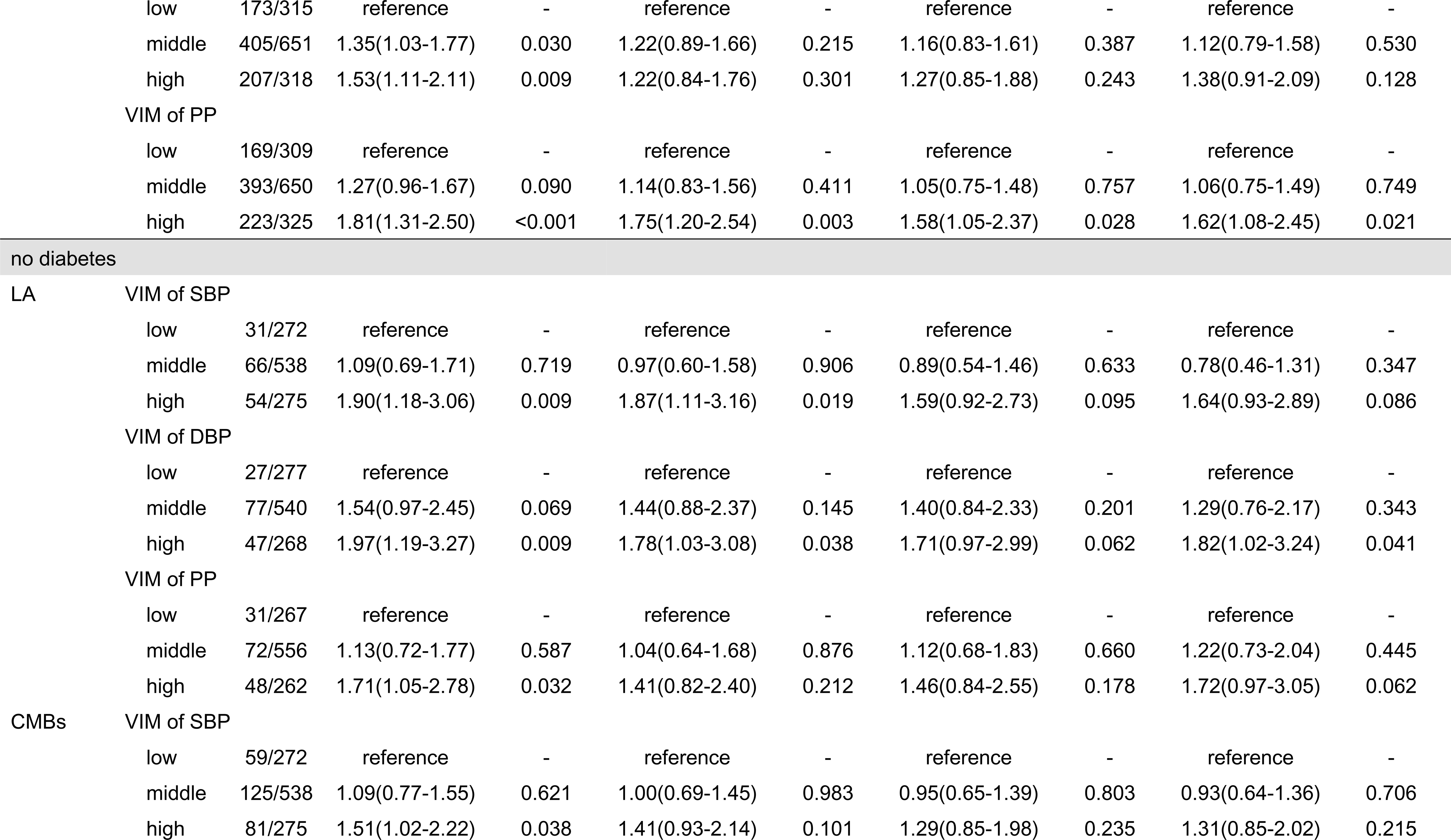
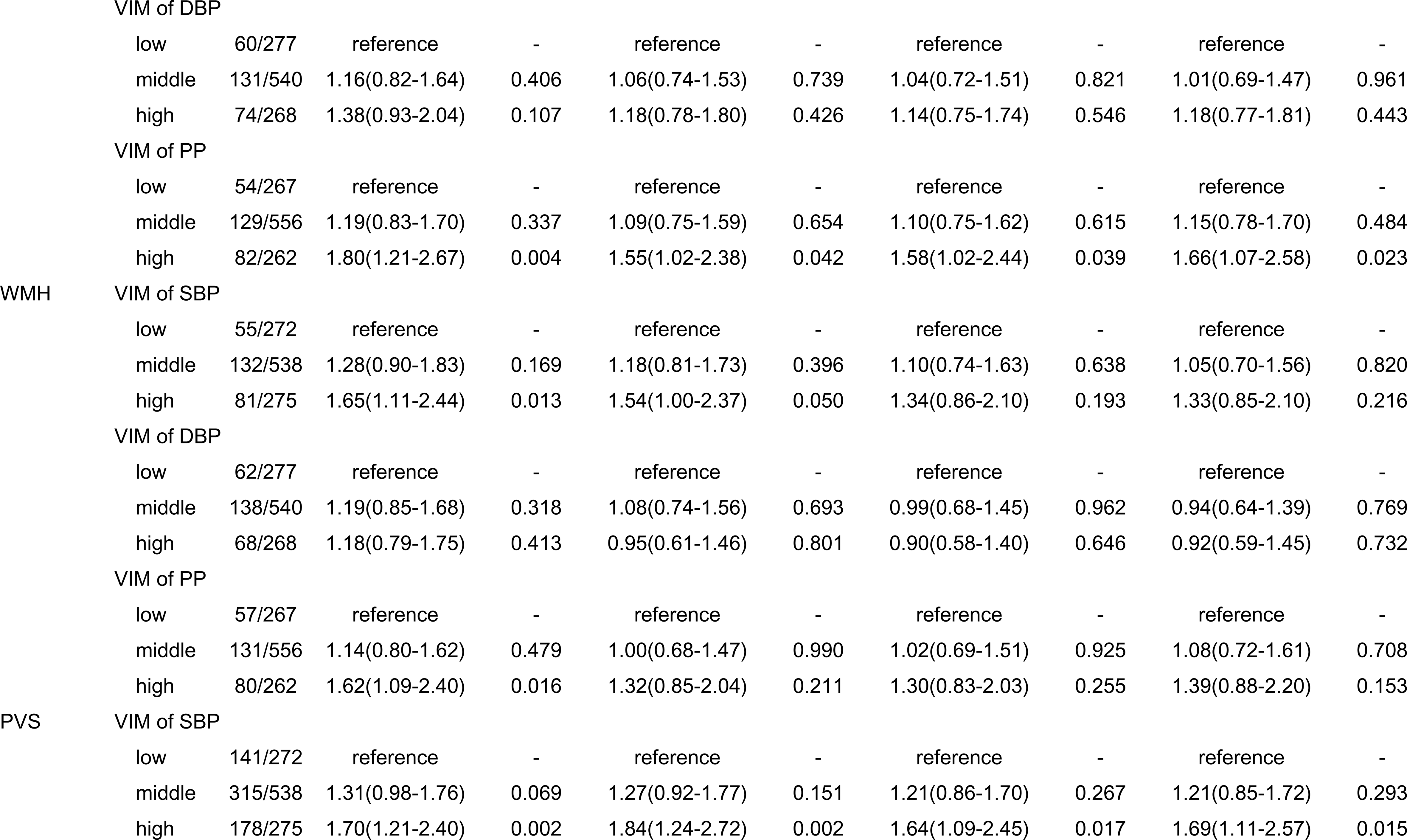
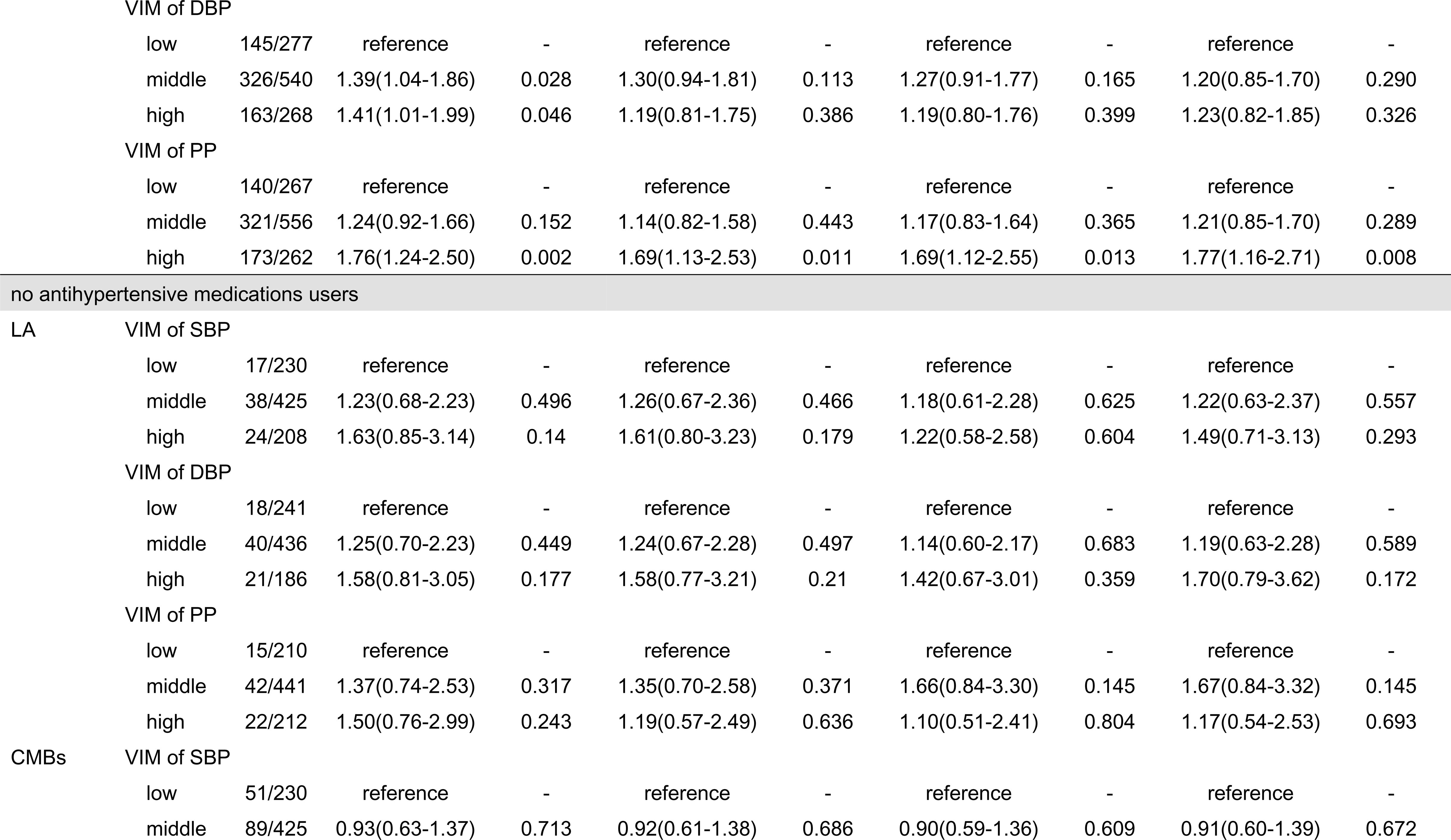
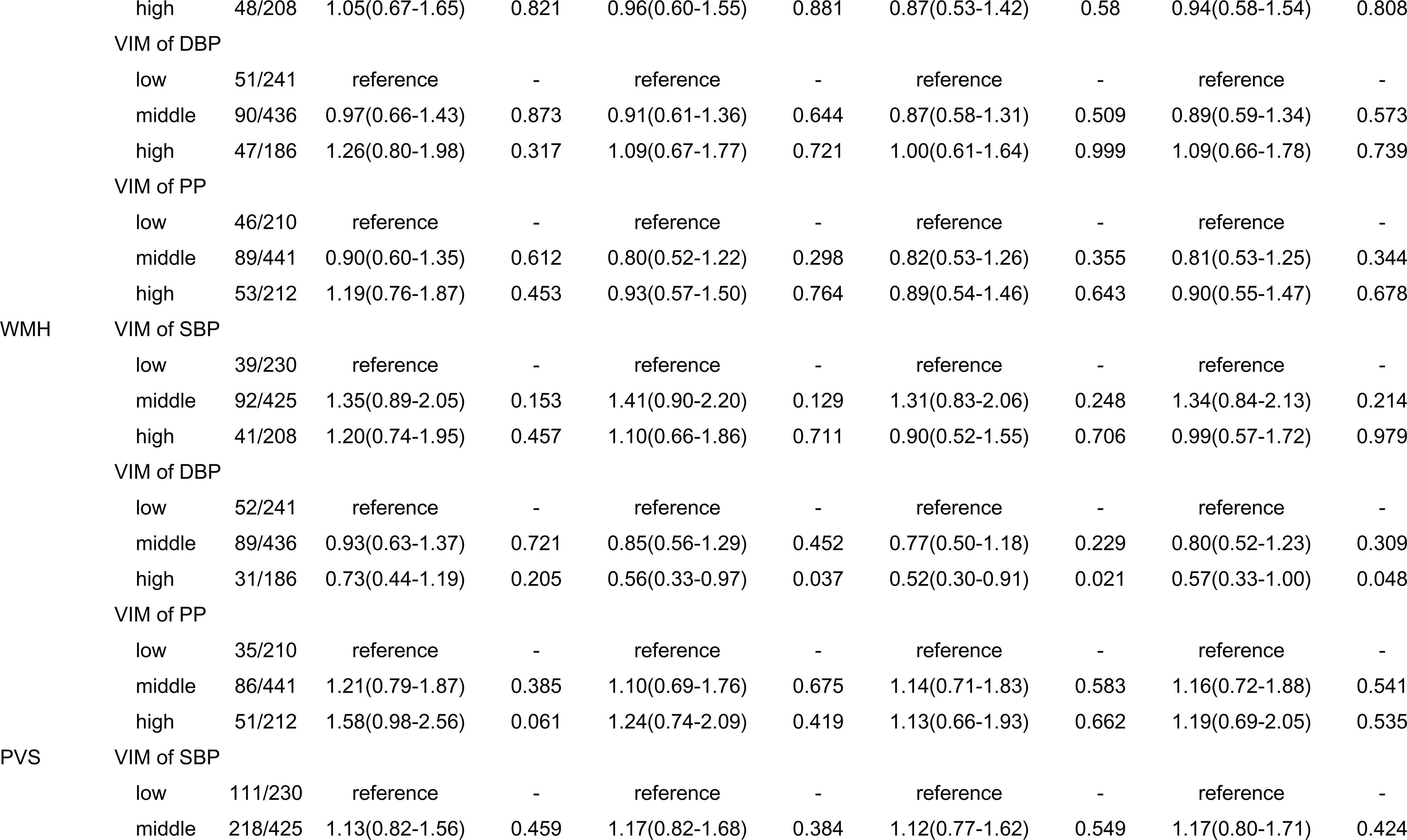
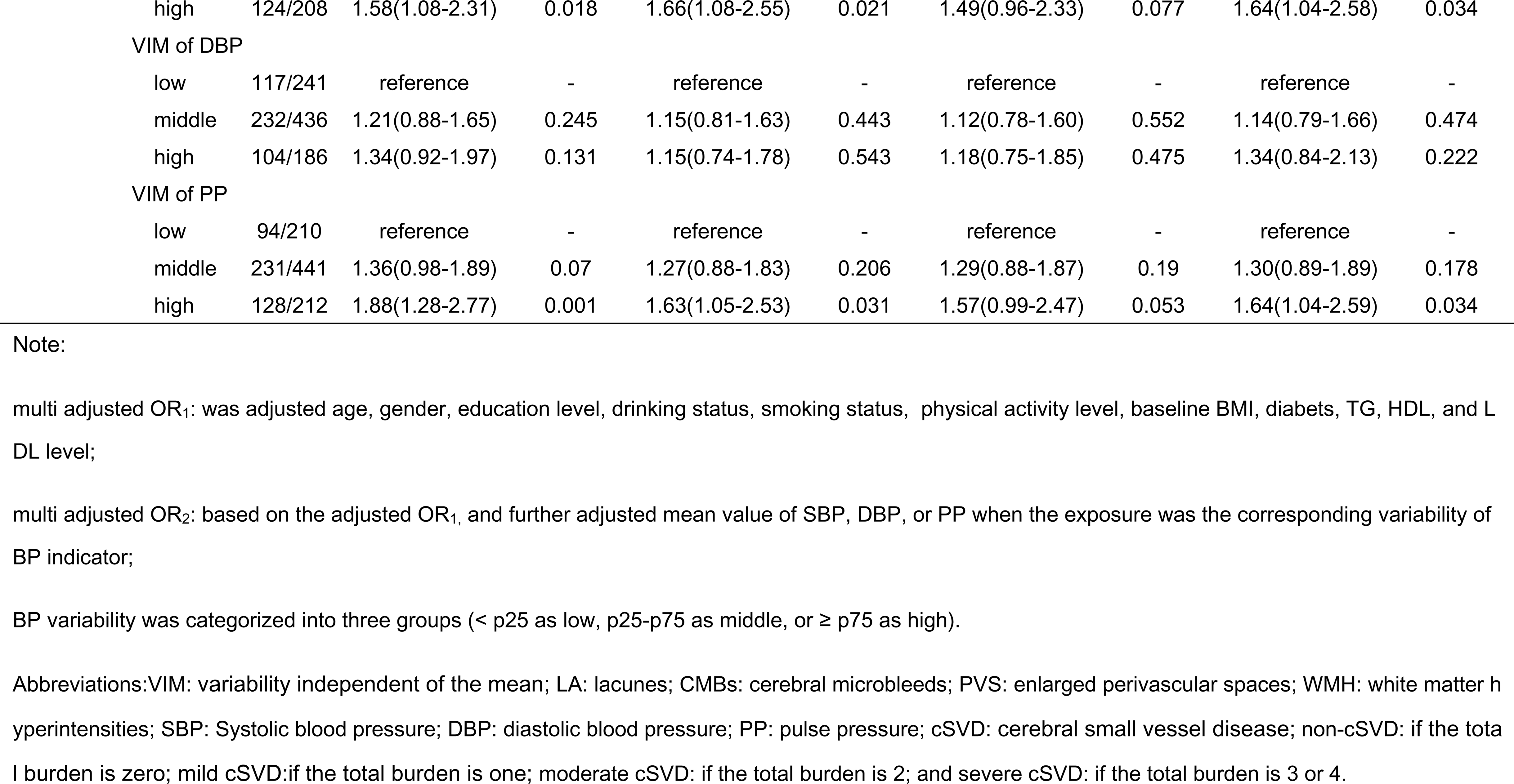
Sensitivity analysis for the estimated association between variability of BP and different markers of cSVD.

